# Molecular Effects of Maternal Cannabis Use on Infant Feeding Regulation

**DOI:** 10.64898/2026.11.23.26368582

**Authors:** Marissa Chow, Natalie Finton, Kiran Singh, Devika Lekshmi, Mario Cordova, Elizabeth Yen

## Abstract

**Background:** Delta-9-tetrahydrocannabinol (THC) regulates appetite through its effect on the hypothalamus, but this is understudied in neonates.

**Objective:** To determine the molecular effects of maternal THC on hypothalamic/reward gene regulation, feeding, and growth in neonates.

**Methods:** Saliva samples of THC-exposed (n=25) and age-matched non-exposed (n=25) neonates underwent transcriptomic analysis of cannabinoid receptors and hypothalamic/reward genes. We explored sex differences and exposure type (THC vs THC+opioids). Continuous data were analyzed with a Student’s t-test and categorical data with a Chi-squared test.

**Results:** THC exposure was associated with upregulation of *CNR1*, *POMC* (p<0.05), and *DRD2* (p=0.06). THC exposure also trended with greater feeding amount and less weight loss in the first 3 days of life. Gene expression also differed by sex and exposure type.

**Conclusion:** Maternal THC use is associated with changes in hypothalamic and reward genes, accompanied by feeding and weight effects.

## INTRODUCTION

Prenatal drug exposure is associated with feeding dysregulation, ranging from poor feeding to hyperphagia. For example, fetal alcohol spectrum disorder is associated with oral aversion and delayed transition to solid foods,^1^ neonatal opioid withdrawal syndrome (NOWS) with excessive yet uncoordinated feeding,^2^ and cocaine exposure with poor and disorganized feeding.^2,3^ Despite the unknown mechanisms, salivary transcriptomic data from our laboratory showed that the effects of opioids on reward signaling may underlie infant feeding dysregulation.^4^ This was evidenced by the upregulation of dopamine receptor 2/*DRD2* (a key reward gene) in opioid-exposed males, which correlated with the amount of milk intake in the first week of life, suggesting that food intake may replace the reward signaling once provided by maternal opioids. Further, maternal opioid use may be associated with hypothalamic imbalance, evidenced by the upregulation of both anorexigenic (leptin receptor/*LEPR*) and reward signaling (*DRD2*).^4^

Expanding our findings in opioid-exposed neonates, we aimed to examine the molecular impact of maternal THC on infant feeding regulation. Since 2002, cannabis use during pregnancy has increased with the legalization of cannabis.^5^ The active ingredient of cannabis, delta-9-tetrahydrocannabinol (THC), provides antiemetic and appetite-inducing properties through its effect on hypothalamic gene expression by binding to cannabinoid receptors (CNR1, CNR2).^6,7^ THC crosses the placenta and is associated with adverse neonatal outcomes, e.g., low birthweight and perinatal mortality.^8,9^ We hypothesized that maternal THC is associated with neonatal dysregulation of the hypothalamic and reward genes via its action on the cannabinoid receptors (primary outcome), and that THC use was linked to greater feeding amount and less weight loss in the first few days of life (secondary outcome). We also explored the effects of infant sex and type of THC (THC-alone vs THC combined with opioids (THC+OPI) on our outcomes.

## METHODS

In this prospective observational pilot study, we enrolled neonates ≥34 weeks’ gestational age (GA) born at Tufts Medical Center between March 2017 and January 2024 to mothers with and without THC use. This study was approved by the Institutional Review Board (IRB). The history of THC and opioid use is based on the diagnosis in electronic medical records, either by maternal urine toxicology, verbal screening, or neonatal urine/meconium toxicology. Neonates with congenital or chromosomal anomalies were excluded. Maternal demographic data included race, ethnicity, delivery method, group B streptococcus/GBS status, cigarette use, and polysubstance use (presence of other psychotropic medications, e.g., anxiolytics, antidepressants, in addition to THC. Neonatal demographics included sex, GA, Apgar at 1 and 5 minutes, anthropometric measurements (birth weight/BW, length/L, head circumference/HC, and percentiles), small for gestational age/SGA status, feeding type (breastmilk, formula, mixed), weight and weight change percentage on day of life (DOL) 2 and 3, and total intake on DOL 2 and 3 (mL/kg/day).

Saliva samples were collected within 48 hours of life using the previously used methods,^10^ Briefly, saliva was collected by gently suctioning between the cheek and the gum for 15-30 seconds (s) using a 1-milliliter (mL) insulin syringe (Becton, Dickinson and Company, Franklin Lakes, NJ) attached to the low-pressure wall suction. Saliva was immediately placed in 250-microliter (µL) RNAprotect Saliva Reagent (Qiagen, Hilden, Germany) to minimize RNA degradation, vortexed, and stored at 4°C pending RNA extraction. Saliva was collected one hour from feeding to minimize milk contamination and before pharmacotherapy to eliminate the effect of postnatal medication(s).

Using the RNeasy Micro Kit (Qiagen), we performed on-column DNase treatment for each sample with RNase-free DNase I Set (Qiagen) to minimize DNA contamination. Samples were then converted to cDNA using SuperSript^TM^ VILO cDNA Synthesis Kit and pre-amplified using TaqMan® PreAmp Master Mix Kit (Applied Biosystems) based upon the inventoried amplicons of the targeted genes of interest, including cannabinoid receptor genes (cannabinoid receptor 1/*CNR1*, cannabinoid receptor 2/*CNR2*), and hypothalamic/reward genes previously studied (dopamine receptor type 2/*DRD2*, neuropeptide Y2 receptor/*NPY2R*, leptin receptor/*LEPR*, and proopiomelanocortin/*POMC*).^4^ Samples were pre-amplified using the Mastercycler® pro S Thermal Cycler (Eppendorf, Hamburg, Germany) at 95⁰ C for 10 minutes (min) for enzyme activation, followed by 14 cycles of pre-amplification at 95⁰ C for 15 s and 60⁰ C for 4 min, then 99⁰ C for 10 min for enzyme inactivation. Pre-amplified cDNA samples were stored at -20⁰ C until qPCR amplification.

Starting total mRNA input across samples was normalized against two reference genes shown to be stable across sex and post-conceptional age, i.e., glyceraldehyde-3-phosphate dehydrogenase (*GAPDH*) and tyrosine 3-mono-oxygenase/tryptophan 5-mono-oxygenase activation protein, zeta polypeptide (*YWHAZ*).^11^ Threshold cycle (Ct) values for genes of interest and reference genes were detected using a QuantStudio^TM^ 7 Flex Real-Time PCR (Applied Biosystems) machine. The following thermal cycle profile was utilized: incubation at 50⁰ C for 2 min, denaturation at 95⁰ C for 10 min, then 40 cycles of denaturation at 95⁰ C for 15 s, and annealing and extension at 60⁰ C for 60 s. All samples were run in duplicate. To ensure plate functionality, water was used as a negative control. A commercial total RNA control (human) (Applied Biosystems^TM^) was used as a positive control and included in every plate to assess plate-to-plate variability. Normalized delta Ct (ΔCt) values were calculated by subtracting the geometric mean Ct values of the reference genes from the mean Ct values of each target gene. ΔCt values are proportional to gene expression levels (more negative values correspond to lower levels).

### Statistical Analysis

We analyzed the data using Stata 17 (StataCorp, College Station, TX), and created figures using GraphPad Prism version 10.5.0 for Windows, GraphPad Software, Boston, Massachusetts, USA (www.graphpad.com). Univariate analyses were performed using a Student’s t-test or Wilcoxon rank-sum test for continuous variables and a Chi-squared test for categorical variables. Multivariate regression was performed for gene expression data (primary outcomes), adjusting for variables that may affect gene expression levels, e.g., SGA, sex, and cigarette smoking.^12–14^ Analysis of variance (ANOVA) was performed to explore the differential effects of THC-alone vs THC+OPI on the outcomes. Finally, to explore sex-specific effects, we analyzed gene expression and feeding data within each sex. All statistical testing was two-tailed with alpha=0.05.

## RESULTS

### Demographic and Clinical Data

We enrolled 25 neonates with prenatal THC exposure and 25 GA-matched (within 1 week) non-exposed neonates. Both cohorts were comparable in sex, race, and ethnicity. THC-exposed neonates were born smaller, had a greater SGA proportion, greater exposure to maternal cigarette use and hepatitis C, and a longer hospital stay (p<0.05, **Table 1**).

**Table 1.**
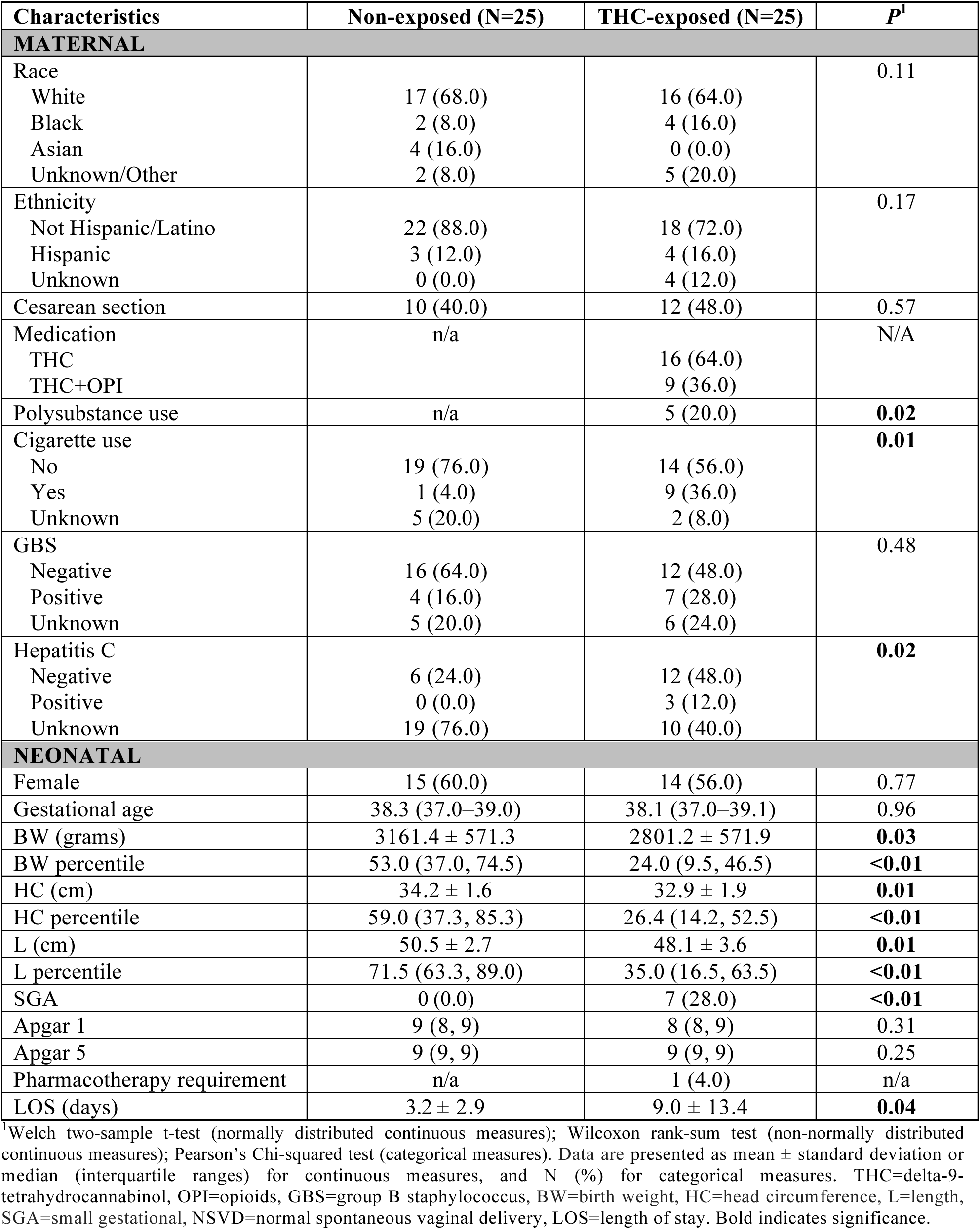
Demographic and Clinical Data.

| Characteristics | Non-exposed (N=25) | THC-exposed (N=25) | *P*<sup>1</sup> |
|---|---|---|---|
| **MATERNAL** |  |  |  |
| Race |  |  | 0.11 |
| White | 17 (68.0) | 16 (64.0) |  |
| Black | 2 (8.0) | 4 (16.0) |  |
| Asian | 4 (16.0) | 0 (0.0) |  |
| Unknown/Other | 2 (8.0) | 5 (20.0) |  |
| Ethnicity |  |  | 0.17 |
| Not Hispanic/Latino | 22 (88.0) | 18 (72.0) |  |
| Hispanic | 3 (12.0) | 4 (16.0) |  |
| Unknown | 0 (0.0) | 4 (12.0) |  |
| Cesarean section | 10 (40.0) | 12 (48.0) | 0.57 |
| Medication | n/a |  | N/A |
| THC |  | 16 (64.0) |  |
| THC+OPI |  | 9 (36.0) |  |
| Polysubstance use | n/a | 5 (20.0) | **0.02** |
| Cigarette use |  |  | **0.01** |
| No | 19 (76.0) | 14 (56.0) |  |
| Yes | 1 (4.0) | 9 (36.0) |  |
| Unknown | 5 (20.0) | 2 (8.0) |  |
| GBS |  |  | 0.48 |
| Negative | 16 (64.0) | 12 (48.0) |  |
| Positive | 4 (16.0) | 7 (28.0) |  |
| Unknown | 5 (20.0) | 6 (24.0) |  |
| Hepatitis C |  |  | **0.02** |
| Negative | 6 (24.0) | 12 (48.0) |  |
| Positive | 0 (0.0) | 3 (12.0) |  |
| Unknown | 19 (76.0) | 10 (40.0) |  |
| **NEONATAL** |  |  |  |
| Female | 15 (60.0) | 14 (56.0) | 0.77 |
| Gestational age | 38.3 (37.0–39.0) | 38.1 (37.0–39.1) | 0.96 |
| BW (grams) | 3161.4 ± 571.3 | 2801.2 ± 571.9 | **0.03** |
| BW percentile | 53.0 (37.0, 74.5) | 24.0 (9.5, 46.5) | **<0.01** |
| HC (cm) | 34.2 ± 1.6 | 32.9 ± 1.9 | **0.01** |
| HC percentile | 59.0 (37.3, 85.3) | 26.4 (14.2, 52.5) | **<0.01** |
| L (cm) | 50.5 ± 2.7 | 48.1 ± 3.6 | **0.01** |
| L percentile | 71.5 (63.3, 89.0) | 35.0 (16.5, 63.5) | **<0.01** |
| SGA | 0 (0.0) | 7 (28.0) | **<0.01** |
| Apgar 1 | 9 (8, 9) | 8 (8, 9) | 0.31 |
| Apgar 5 | 9 (9, 9) | 9 (9, 9) | 0.25 |
| Pharmacotherapy requirement | n/a | 1 (4.0) | n/a |
| LOS (days) | 3.2 ± 2.9 | 9.0 ± 13.4 | **0.04** |

<sup>1</sup>Welch two-sample t-test (normally distributed continuous measures); Wilcoxon rank-sum test (non-normally distributed continuous measures); Pearson’s Chi-squared test (categorical measures). Data are presented as mean ± standard deviation or median (interquartile ranges) for continuous measures, and N (%) for categorical measures. THC=delta-9-tetrahydrocannabinol, OPI=opioids, GBS=group B staphylococcus, BW=birth weight, HC=head circumference, L=length, SGA=small gestational, NSVD=normal spontaneous vaginal delivery, LOS=length of stay. Bold indicates significance.

### Primary Outcome: Gene Expression Data

**Figure 1** demonstrates greater expression of *CNR1* and *POMC* in THC-exposed than non-exposed neonates (ΔCt mean ± SD: -8.7 ± 4.7 vs -11.3 ± 4.3 for *CNR1*, p=0.043; -12.2 ± 4.8 vs - 15.2 ± 3.2 for *POMC*, p=0.015). *DRD2* expression also trended higher in neonates with THC exposure (-12.6 ± 3.4 vs -14.3 ± 3.0, p=0.07). After adjusting for variables that may modify gene expression, e.g., SGA, sex, and cigarette smoking,^12–14^ the upregulation of *CNR1* and *POMC* in THC-exposed neonates remained significant (ΔCt β ± SE: 2.57 ± 1.56 for *CNR1*; 3.48 ± 1.41 for *POMC*, p<0.05).

**Figure 1.**
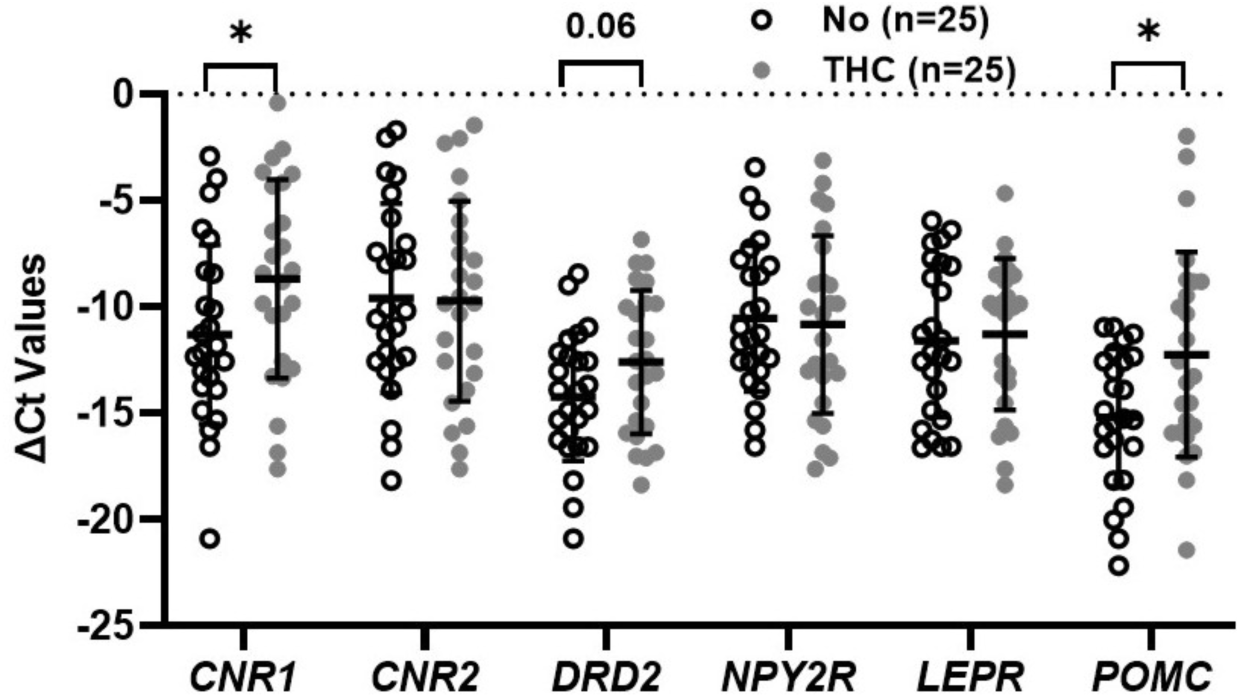
Gene Expression Changes by THC Exposure. THC exposure may create an imbalance between hedonic/reward and anorexigenic signaling, as shown by the upward trend in *DRD2* expression and significantly higher POMC expression. These changes may be mediated by THC effects on its receptor, evidenced by significantly greater expression of *CNR1*. *CNR1*: cannabinoid receptor type 1, *DRD2*: dopamine receptor type 2, *NPY2R*: neuropeptide Y2 receptor, *LEPR*: leptin receptor, *POMC*: proopiomelanocortin, ΔCt: delta cycle threshold, more negative numbers indicate lower gene expression, THC: tetrahydrocannabinol.

### Secondary Outcomes: Feeding and Weight Data

Because hospital stay varied, we evaluated feeding in the first 3 days of life, when most neonates were still hospitalized. Feeding amount and percent weight loss on DOL 2 and 3 did not differ significantly between the THC- and non-exposed cohorts (**Figure 2**). Despite the non-significant differences, THC-exposed neonates had less weight loss and higher feeding volume in the first 3 days of life. THC exposure also trended with less breast milk use (48.0% vs 75.0% on DOL 2, p=0.06; 31.6% vs 72.7% on DOL3, p=0.09).

**Figure 2.**
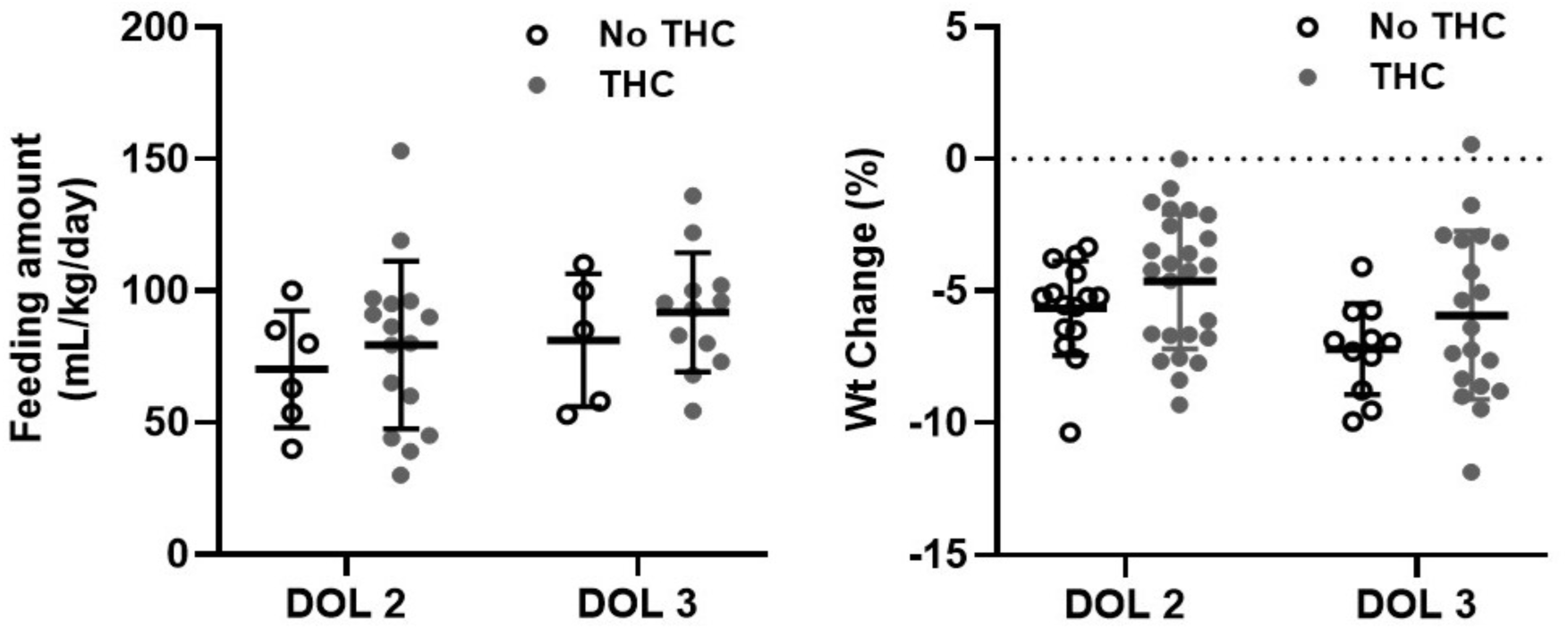
Feeding Amount and Weight Change by THC Exposure. THC-exposed infants had slightly higher feeding amounts (left panel) and less weight loss on days 2 and 3 (right panel). DOL: day of life, THC: tetrahydrocannabinol.

### Exploratory Outcome: Sex-specific Effects of THC Exposure

Compared to no exposure, THC exposure was associated with upregulation of *POMC* in females and *DRD2* in males (**Figures 3A, B**). Feeding amounts on DOL 2 and 3 did not differ significantly between sexes (**Figures 3C, E**). Females with THC exposure lost less weight on DOL 2 compared to non-exposed females (**Figure 3D**); otherwise, there was no significant difference in weight loss for both sexes on DOL 2 and 3 (**Figures 3D, F**).

**Figure 3.**
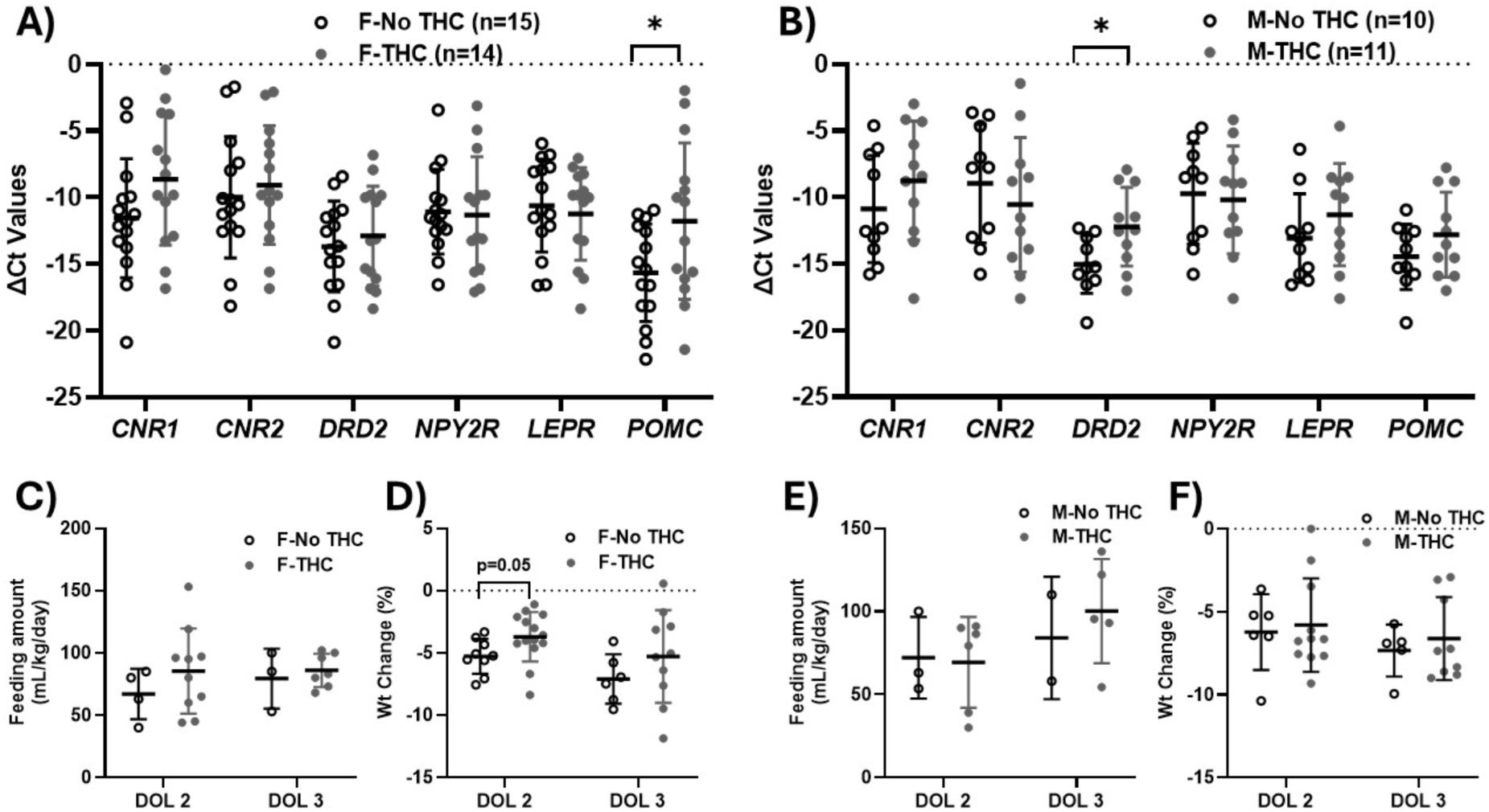
Sex-specific Changes Related to THC Exposure. THC exposure is associated with greater anorexigenic signaling in females, evidenced by significantly greater *POMC* expression (**A**), whereas in males, it is associated with greater reward/hedonic signaling, evidenced by significantly greater *DRD2* expression (**B**). THC exposure trended with greater feeding amounts and less weight loss in both females (**C, D**) and males (**E, F**), with significantly less weight loss in THC-exposed females on DOL 2 (**D**). The clinical findings in THC-exposed females, along with the greater expression of *POMC*, may suggest the paradoxical effect of THC on this anorexigenic gene, which has been shown in animal studies.^21,22^ *CNR1*: cannabinoid receptor type 1, *DRD2*: dopamine receptor type 2, *NPY2R*: neuropeptide Y2 receptor, *LEPR*: leptin receptor, *POMC*: proopiomelanocortin, ΔCt: delta cycle threshold, more negative numbers indicate lower gene expression, THC: tetrahydrocannabinol.

### Exploratory Outcome: Effects by Type of Exposure (THC-alone vs THC+OPI)

As shown in **Table 2**, the expression of *POMC* differed significantly across cohorts, with the highest expression in the THC+OPI cohort. *CNR1* expression also trended higher in the THC-alone cohort, followed by the THC+OPI cohort, and was lowest in the non-exposed cohort. Neonates exposed to THC+OPI were born the smallest, had the highest SGA proportion, and the longest hospital stay compared to those in the THC-alone or non-exposed cohorts. THC+OPI cohort had the highest maternal cigarette smoking and hepatitis C positivity. Compared to the THC-alone cohort, those in the THC+OPI cohort also had greater polysubstance exposure. Feeding and weight data did not differ significantly across the cohorts, but neonates exposed to THC-alone had higher volume intake and lost less weight on both DOL 2 and 3.

**Table 2.**
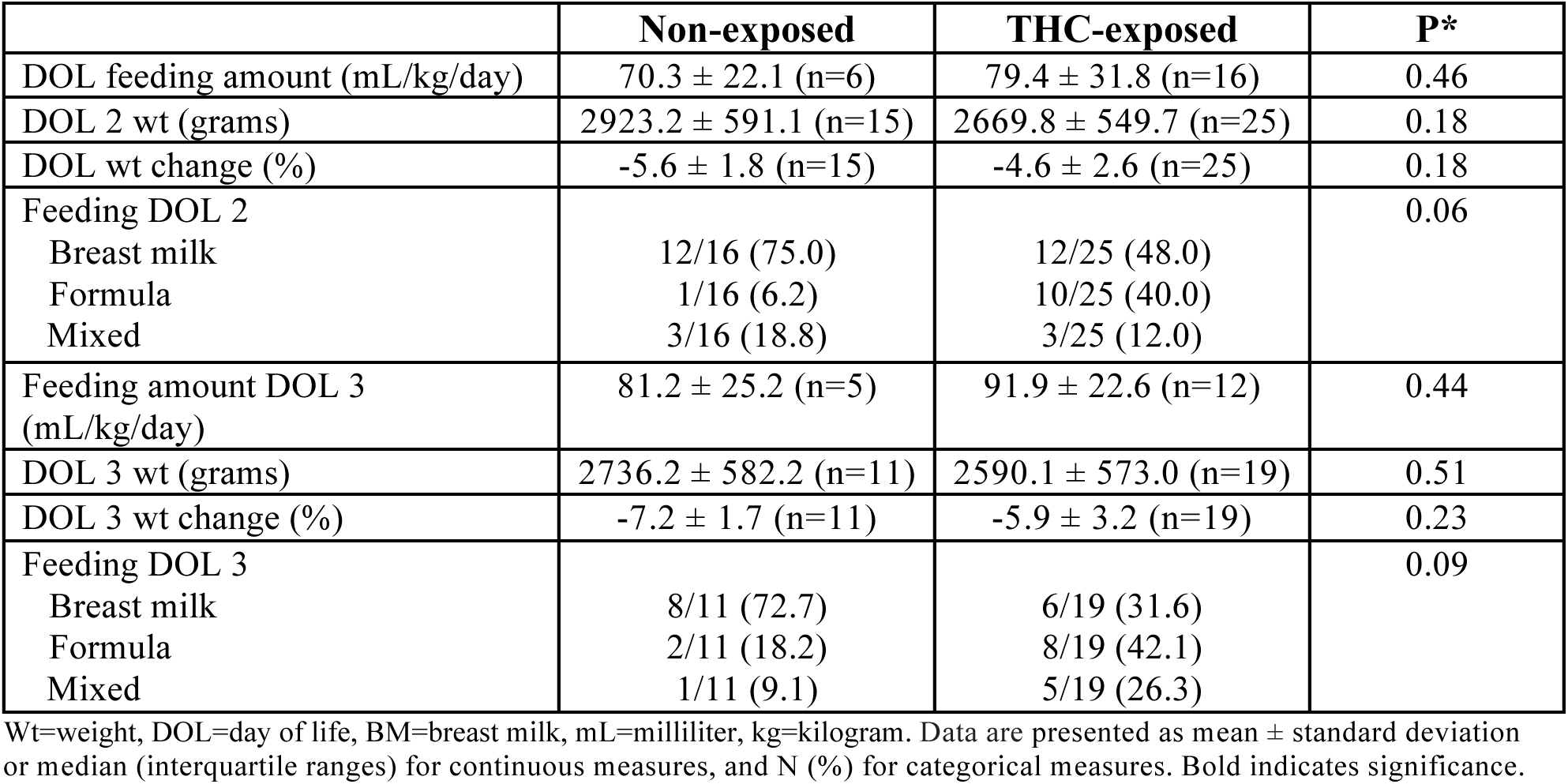
Weight and Feeding Data by Exposure.

**Table 3.** Demographic and Clinical Data by Types of THC.

| **Variables** | **No THC<br>(n=25)** | **THC-Alone<br>(n=16)** | **THC+OPI<br>(n=9)** | **p-value** |
|---|---|---|---|---|
| **MATERNAL** |  |  |  |  |
| Race |  |  |  | 0.093 |
| White | 68% (17) | 56% (9) | 78% (7) |  |
| Black | 8% (2) | 12% (2) | 22% (2) |  |
| Asian | 16% (4) | 0% (0) | 0% (0) |  |
| Unknown/Other | 8% (2) | 31% (5) | 0% (0) |  |
| Ethnicity |  |  |  | **0.041** |
| Not Hispanic/Latino | 88% (22) | 75% (12) | 67% (6) |  |
| Hispanic/Latino | 12% (3) | 6% (1) | 33% (3) |  |
| Unknown | 0% (0) | 19% (3) | 0% (0) |  |
| Cesarean section | 40% (10) | 50% (8) | 44% (4) | 0.82 |
| Polysubstance use | NA | 6% (1) | 44% (4) | **0.022** |
| Cigarette use |  |  |  | **<0.001** |
| No | 76% (19) | 81% (13) | 11% (1) |  |
| Yes | 4% (1) | 12% (2) | 78% (7) |  |
| Unknown | 20% (5) | 6% (1) | 11% (1) |  |
| GBS |  |  |  | 0.76 |
| Negative | 64% (16) | 44% (7) | 56% (5) |  |
| Positive | 16% (4) | 31% (5) | 22% (2) |  |
| Unknown | 20% (5) | 25% (4) | 22% (2) |  |
| Hep C |  |  |  | **0.024** |
| Negative | 24% (6) | 44% (7) | 56% (5) |  |
| Positive | 0% (0) | 6% (1) | 22% (2) |  |
| Unknown | 76% (19) | 50% (8) | 22% (2) |  |
| **NEONATAL** |  |  |  |  |
| Female | 60% (15) | 44% (7) | 78% (7) | 0.24 |
| GA (weeks) | 38.3 (37.0-39.0) | 38.0 (37.0-39.1) | 38.2 (37.1-39.2) | 0.99 |
| BW (g) | 3161.4 (571.3) | 2901.9 (638.6) | 2622.1 (400.3) | **0.05** |
| BW percentile | 53.5 (22.6) | 36.5 (28.9) | 21.6 (14.7) | **0.003** |
| Length (cm) | 50.5 (2.7) | 48.9 (3.5) | 46.5 (3.5) | **0.009** |
| Length percentile | 71.5 (63.5-89.0) | 43.0 (26.5-85.0) | 19.0 (8.0-36.0) | **<0.001** |
| HC (cm) | 34.2 (1.6) | 33.0 (2.1) | 32.6 (1.4) | **0.04** |
| HC percentile | 59.0 (38.5-84.5) | 26.4 (13.0-67.0) | 33.0 (17.0-36.0) | **0.003** |
| SGA | 0% (0) | 25% (4) | 33% (3) | **0.01** |
| Apgar score (1 min) | 9.0 (8.0-9.0) | 8.0 (8.0-9.0) | 8.0 (7.0-8.0) | 0.21 |
| Apgar score (5 min) | 9.0 (9.0-9.0) | 9.0 (9.0-9.0) | 9.0 (9.0-9.0) | 0.37 |
| Length of stay (days) | 2.0 (2.0-3.0) | 3.0 (2.0-5.0) | 6.0 (5.0-14.0) | **0.005** |
| **FEEDING/WEIGHT DATA** |  |  |  |  |
| DOL 2 feeding amt (ml/kg/day) | 71.6 (20.1) | 86.3 (32.9) | 70.5 (30.5) | 0.50 |
| DOL 2 wt (grams) | 2884.3 (559.0) | 2772.2 (611.4) | 2487.7 (383.8) | 0.22 |
| DOL 2 wt change (%) | -5.7 (1.8) | -4.4 (2.5) | -5.1 (2.8) | 0.31 |
| DOL 3 feeding amt (ml/kg/day) | 81.2 (25.2) | 100.6 (18.3) | 83.2 (24.6) | 0.31 |
| DOL 3 wt (grams) | 2736.2 (582.2) | 2723.6 (688.1) | 2441.7 (398.9) | 0.46 |
| DOL 3 wt change (%) | -7.2 (1.7) | -5.0 (3.2) | -7.0 (2.9) | 0.13 |
| Feeding type DOL 2 |  |  |  | 0.07 |
| Breast milk | 68% (13) | 62% (10) | 22% (2) |  |
| Formula | 11% (2) | 31% (5) | 56% (5) |  |
| Mixed | 21% (4) | 6% (1) | 22% (2) |  |
| Feeding type DOL 3 |  |  |  | 0.11 |
| Breast milk | 73% (8) | 50% (5) | 11% (1) |  |
| Formula | 18% (2) | 30% (3) | 56% (5) |  |
| Mixed | 9% (1) | 20% (2) | 33% (3) |  |

THC: tetrahydrocannabinol, OPI: opioids, Hep: hepatitis, BW: birth weight, HC: head circumference, SGA: small for gestational age, amt: amount, DOL: day of life, Wt: weight, mL: milliliter, kg: kilograms. Data are presented as mean ± standard deviation or median (interquartile ranges) for continuous measures, and N (%) for categorical measures. Bold indicates significance.

## DISCUSSION

Using neonatal saliva, we demonstrated differential expression of genes involved in cannabis and feeding-regulation pathways that may relate to feeding behavior and growth in the immediate postnatal period.

### Demographic and Clinical Data

Supporting previous studies, THC-exposed neonates in our study were born significantly smaller and with a higher incidence of SGA.^15^ A greater proportion of non-exposed infants were fed breast milk compared to THC-exposed infants, which may reflect hospital-wide practice adherent to the American Academy of Pediatrics recommendations, discouraging the use of breast milk and breastfeeding in mothers with THC use.^16^

### Primary and Secondary Outcomes: THC Exposure and Association with Transcriptomic, Feeding, and Weight Changes

The upregulation of *CNR1* in the THC-exposed cohort confirms the biological effects of THC by acting on this receptor. The upregulation of *POMC* and *DRD2* in the THC-exposed cohort suggests that THC may modulate hypothalamic and reward genes known to regulate feeding behavior, similar to the effects of opioids.^4^ While non-significant, the greater feeding amount in the THC-exposed cohort on both DOL 2 and 3 supports adult and animal data demonstrating the crucial role of *CNR1* in stimulating appetite/hunger and increasing the drive to ingest high-fat/high-sugar diets.^17,18^ Its role in appetite regulation is among the reasons for increased cannabis use in pregnancy in recent decades.^19^ Not only is *CNR1* important in adults, but this gene is also fundamental even in infancy by triggering suckling behavior. Animal studies showed that *CNRI*-knockout pups lacked milk suckling, which affected their survival rate.^20,21^ Paralleling the greater feeding amount in the THC-exposed neonates, these neonates also experienced less weight loss in the first 3 days of life. Although not significant, these results may suggest that maternal THC use may affect offspring feeding behavior and growth. We will validate these findings using a larger sample size to elucidate the impact of maternal THC use on offspring growth, nutrition, and health outcomes.

The upregulation of both *POMC* (anorexigenic signaling) and *DRD2* (hedonic/reward signaling) in THC-exposed neonates resembles our prior study showing the upregulation of *LEPR* and *DRD2* in opioid-exposed neonates.^4^ Activation of *POMC* leads to satiety, and its deficiency can lead to early-onset obesity, adrenal insufficiency, and hyperphagia.^22^ Activation of *DRD2,* on the other hand, promotes reward signaling and induces feeding behaviors in pups presented with an artificial nipple compared to their non-manipulated counterparts.^22,23^ Therefore, considering the higher feeding amount in THC-exposed neonates in our study, our findings suggest that THC exposure may be associated with an imbalance in these two opposing effects. Koch et al. demonstrated that cannabinoid receptor activation induces feeding in mice. However, contrary to their hypothesis, neuronal activity of POMC cells in these sated mice was upregulated rather than downregulated.^24^ Further, recent data also demonstrated that activation of POMC neurons stimulated sugar consumption in the fed state via activation of opioid β-endorphin.^25^ Such a paradoxical effect highlighted the unique role of POMC neurons in cannabis-related feeding behavior, which may predispose to overconsumption. A combined human-animal study in the future may help elucidate this paradoxical effect of maternal THC use on infant reward and satiety signaling and long-term metabolic health.

### Sex-specific Effects on Gene Expression, Feeding, and Weight Changes

Our study shows distinct sex-specific effects of THC via the upregulation of *POMC* in females with THC exposure and *DRD2* in males with THC exposure. This molecular effect is paralleled by greater feeding amount and less weight loss on DOL 2 and 3 in THC-exposed males and females compared to their non-exposed counterparts. The effect of THC on the *POMC* pathway in females supports animal studies^26^ showing greater activation of *POMC* neurons in females in response to cannabis. Given animal data showing the paradoxical effect of cannabis on POMC activation and overconsumption of sugary foods despite being satiated,^24,25^ future studies should focus on examining THC-related upregulation of *POMC* in females and effects on feeding behavior and risk of obesity. Additionally, pending further validation, sex-specific changes in gene expression could inform distinct interventions targeting the *POMC* versus *DRD2* pathways.

### Type of THC Exposure and Association with Outcomes

Within the THC-exposed neonates, we stratified by exposure (THC-alone vs THC+OPI) to explore if the combination of opioids with THC was associated with molecular and behavioral effects compared to THC-only exposure. Current data suggest that THC+OPI was linked to worse clinical outcomes (smaller size at birth, greater SGA incidence, and longer hospital stay). On the molecular level, the gene expression levels were similar between THC-alone and THC+OPI cohorts, indicating that co-exposure with opioids may not have synergistic or deletory effects on the hypothalamic and reward genes. Compared to no exposure, THC+OPI may have a different action compared to THC-alone, with THC-alone exposure linked to significant upregulation in *CNR1* and *POMC* expression, while THC+OPI trended toward greater expression of *DRD2* and some *POMC*. Feeding and weight data did not show significant differences between THC-alone and THC+OPI exposure, with a trend of greater feeding amount and less weight loss in the THC-alone cohort on DOL 2 and 3. Whether this trend is associated with the significant upregulation of *CNR1* and *POMC* in the THC-alone cohort needs to be further validated.

The interaction between THC and opioids and its effect on feeding dysregulation has not been explored, but it has been studied in the context of analgesic effects. Preclinical and clinical data demonstrated that THC reduced opioid doses for analgesia and the severity of opioid withdrawal, suggesting THC may have an opioid-sparing effect.^27^ However, a recent randomized placebo-controlled trial did not show synergistic analgesic effects between THC and opioids in adults with knee osteoarthritis.^28^ Rather, the combination of cannabinoids and opioids was linked to impaired working memory reaction time. These findings demonstrate the complexity of the interaction between THC and opioid use, hence an area that warrants future investigation.

### Strength and Limitations

Strengths of our study include non-invasive methods of data collection to assess the effects of maternal THC use on infant outcomes. Saliva collection and feeding/growth measurements posed little to no risk to infants and mothers. Research on the effects of THC exposure has focused on birth outcomes (prematurity, stillbirth, small size),^15^ cognitive and neurobehavioral outcomes,^29,30^ and metabolic health.^31^ Our study provides a novel focus by studying the impact of maternal opioid use on genes regulating feeding behavior and association with postnatal growth, therefore, adding to the body of knowledge on this understudied, yet critical, topic. Further, the sex-specific effects reported in this study also support the importance of studying sex as a biological variable. Finally, while exploratory, the differential gene expression between THC-alone and THC+OPI may serve as a foundation to study the long-term effects of THC exposure in conjunction with other co-exposure. Overall, our study provides important findings that highlight the necessity of monitoring the long-term outcomes following in utero THC exposure.

Limitations of our study include a small sample size and missing data. With only 50 neonates, our study’s generalizability and statistical power are limited. Thus, findings should be interpreted with caution due to the risk of inflated type I error, as we could not correct for multiple comparisons. Given that most neonates were discharged in the first 2-3 days of life, data beyond this period became scarce, making it challenging to compare outcomes with neonates who stayed longer, especially the THC+OPI cohort. Feeding amount data were not always available, either because records were missing or because infants were breastfed. Additionally, our study was limited to hospital stays, restricting our ability to assess long-term growth or developmental outcomes. Lastly, while we stratified our data by the type of THC (alone or in combination with opioids), we did not account for many characteristics of this medication, e.g., dose, route of ingestion, duration of use, which are likely to affect our outcomes.

## CONCLUSION

Our study demonstrates the molecular and behavioral effects of maternal THC use on infant feeding and growth in the immediate period after birth. While limited by the small sample size, data in this study provide evidence suggesting aberrant feeding dysregulation following in utero THC exposure, which may be mediated by an imbalance between the anorexigenic and hedonic/reward signaling. Further, THC exposure may affect males and females differently, predisposing these neonates to sex-specific health problems in adulthood. Our results need to be validated using a larger sample size and should focus on the long-term effects of THC exposure, including metabolic and neurodevelopmental measurements. Despite being non-significant, the trend of greater feeding amount and less weight loss in the THC-exposed cohort in the first few days of life may set these neonates on a growth trajectory with unclear metabolic risks.

## Data Availability

All data produced in the present study are available upon reasonable request to the authors.

